# Where will they grow from here? Association between height at 24 months and adult attained height in a population with a high prevalence of linear growth restriction

**DOI:** 10.1101/2020.08.20.20178905

**Authors:** Julia Schwarz, Pablo Peñataro Yori, William K. Pan, Maribel Paredes Olortegui, Robert Klapheke, Margaret N. Kosek

## Abstract

This was a longitudinal observational cohort study to investigate the association of stunting at 24 months of age and attained adult height. A cohort of 104 Peruvian males (n = 47) and females (n = 57) was studied from 24 months to after puberty in a rural community in the Peruvian Amazon located 15 km southeast from the urban center of Iquitos. Anthropometric measures were made at 24 months and after puberty. Height for Age Z (HAZ) Scores were calculated and compared to assess the association of HAZ score at 24 months and adult height. 67.7% of males and 54.1% of females who were stunted at 24 m (HAZ<-2.0) recovered (HAZ>-2.0) after puberty, whereas 6.25% of males and 6.06% of females who were not stunted at 24 m (HAZ>-2.0) became stunted (HAZ<-2.0) after puberty. The first 1000 days are not the only critical window for growth promotion. Global and national initiatives to reduce stunting should not exclusively focus on the first two years of life, but also work to promote the nutritional initiatives to promote optimal nutrition and growth throughout childhood and adolescence.

## Introduction

Linear growth is a key measure of child health. A link between stunting and both physical and cognitive damage has been demonstrated in the last several decades with increasing validation and stunting is a commonly accepted surrogate for loss of human potential at the population level.^1, 2, 3, 4, 5^ In previous studies, 1 cm gained in the cohort’s average height was associated with a 6% increase in income per capita.^6^

Stunting is now considered a major global health priority and is the focus of several high‐profile initiatives, including Scaling Up Nutrition, the Zero Hunger Challenge and the Nutrition for Growth Summit.^7^ It is estimated that in 2010, 26.7% (95% CI 24.8,28.7) of children or 171 million (167 million in developing countries) were stunted, meaning they had a height-for-age- Z-Score (HAZ) <-2.^8^ For the last decade, growth faltering research has focused on the first 1000 days of life as the window of opportunity for growth promotion.^9^ These studies led to the creation of programs that specifically promote nutrition during the first 24 months. However, recent longitudinal cohort studies have suggested that recovery from growth stunting is possible after the first two years of life.^10, 11, 12^

This study investigates the veracity of the belief that stunting in the first two years dictates height throughout adulthood. Human growth velocity is highly age dependent and has classically been described as occurring in the infancy phase, which begins in midterm gestation and ends at 3–4 years with a pattern of rapid deceleration, the childhood phase of slowly decelerating linear growth velocity, and the pubertal phase, which is characterized by an increase in growth velocity followed by a rapid and steep decline after puberty growth when the epiphyseal plate closes and linear growth ends.^13, 14^ The biology of the childhood and pubertal phases is not accounted for in the many public health programs designed with primary outcomes based on measurements taken at 2 and 5 years. The evidence presented here offers an uncommon perspective in that it follows a cohort from early childhood to puberty and concentrates on HAZ scores at the individual level. Here we analyzed the attained height of children at 12–16 years old who were and were not stunted at 24 months to evaluate the predictive capacity of early childhood measures on final attained adult stature.

## Methods

### Study setting

Santa Clara is a rural community in the Peruvian Amazon located 15 km southeast from the urban center of Iquitos. Stunting in this community is high, whereas acute undernutrition is relatively uncommon. Previous studies offer additional details of this community.^15^

### Study design

Between October 1, 2002 and April 15, 2006, the team conducted a 43-month long prospective, community-based diarrheal disease surveillance in 442 children < 72 months of age in Santa Clara. The cohort and study design have been previously described.^16, 17^ The study cohort was chosen from a community census that generated a list of children < 72 months of age. Enrollment was limited to one child per household. Selection included every third child on the census list sorted according to date of birth. Ninety-six percent of families invited to participate gave informed consent for enrollment in the study protocol. During the months they were enrolled in the study subject’s height was measured monthly by using a marked platform with a sliding footboard. After 72 months of age, the child was removed from surveillance and the next live birth in that community was enrolled.

Follow-up measurements were conducted between September and December 2016. Contact was attempted with all participants in the original cohort. Community health workers returned to the households they had visited monthly during the first stage of the cohort study to see if participants would consent to further measurements. Retention is detailed in Figure 1. Subjects had their height and weight measured and age of puberty was assessed by maternal report of the onset of menses (for female subjects) or the age of voice change (for male subjects). Menarche is an established indicator as the final stage of female puberty^18^ and age of voice change has been shown to be representative of physical maturity in men.^19, 20^ The age of pubertal onset is an important indicator of adult height, as the release of gonadal steroids (estrogen, testosterone) causes closure of the epiphyseal growth plates and the end of statural growth. Therefore, we included only those in the cohort that had gone through puberty as a marker of having passed through the late adolescent growth spurt and be at, or very close to, terminal adult height.

**Figure 1:**
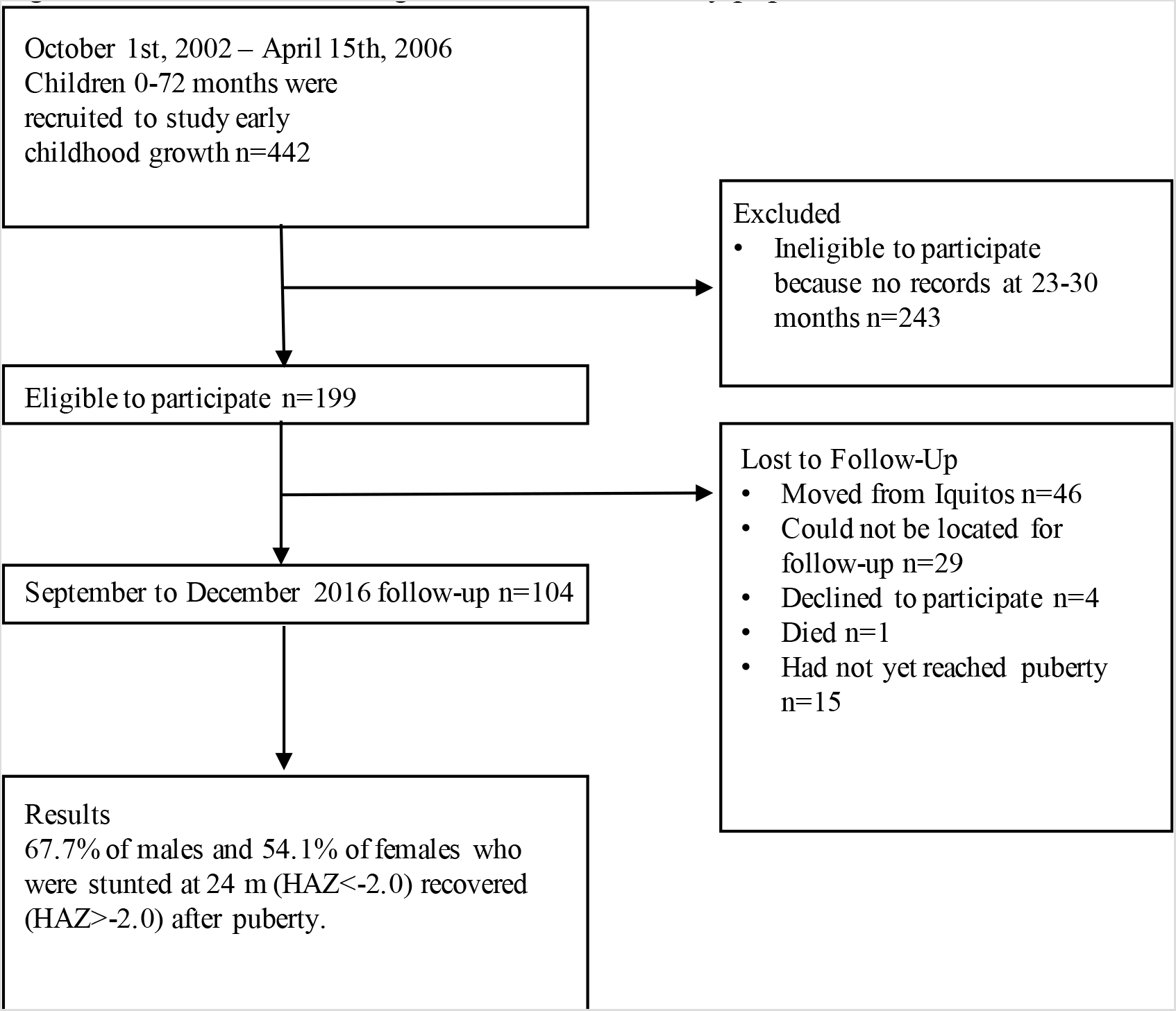
Flow chart showing identification of study population.

### Statistical Analysis

Data was analyzed to look at the association of height at 24 months on height, weight and BMI post-puberty as well as on the age of puberty onset.

The anthropometrical measurements closest to 24 months (ranging from 23–30 months) was compared with the WHO Reference Data 2006 to calculate the HAZ score for each child at 24 months. Participants were then categorized as stunted (HAZ < –2) or not stunted (HAZ>-2) at 24 months based on the HAZ calculated for the date closest to 24 months. For those retained at follow-up, HAZ and BMI post puberty were calculated. The participants were again categorized into “HAZ < –2 post-puberty” and “HAZ > –2 post puberty” based on the anthropometrical measurements after maternal report of puberty.

Based on the change in HAZ score from 24 months to post puberty, participants were divided into four categories. Children who were stunted at 24 months were categorized as “recovered children” if their post-puberty HAZ was greater than negative two, or as “stunted throughout” if their post-puberty HAZ was less than negative two. Among “recovered children,” a subcategory of “fully recovered” was created for those with a post puberty HAZ greater than negative one. Children who were not stunted at 24 months were categorized either as “late stunting” if their post puberty HAZ was less than negative two, or “non-stunted throughout” if their post-puberty HAZ was greater than negative two. The percentage of male, female and total cohort members in each category was calculated. A chi-squared test was used to compare the percentage of male and female participants in each category. In addition, mean attained height among male and female participants was compared across each of the four categories. A one-way ANOVA was conducted to compare the average age of puberty between each category followed by post-hoc comparison using a Tukey test.

Selection bias due to loss-to-follow up was evaluated by comparing sex, HAZ score, and stunting at 24 months among children lost to follow-up and those retained. In addition, using data from 2002–2006, when the study began, average years of maternal education and average monthly income was compared among children lost to follow-up and those retained. Data was missing for 5 members in the group who were measured for adult-follow and 13 members who were lost-to follow up. Two sample t-tests were used to compare means and chi-squared tests compared percentages.

Post-puberty HAZ, weight and BMI were compared across three levels of HAZ at 24 months: HAZ less than negative two, HAZ between negative one and negative two, and HAZ greater than –1. A one –way ANOVA was conducted to compare the association of HAZ at 24 months on HAZ, BMI, and weight at puberty. This was followed by post-hoc comparison using a Tukey test.

Odds ratios were calculated for the overall likelihood a child with a HAZ less than negative two at 24 months will be stunted post puberty compared with a child with a HAZ greater than negative two at 24 months.

The average age of puberty onset was calculated and analyzed for each category. Because puberty onset was reported in years, when puberty onset was studied in months it would have been falsely clustered in the first month of the year. To give a more accurate representation, a random normally distributed covariate with a mean of 6 and standard deviation of 4 was added to the reported age of puberty onset in months.

### Ethical Review

The study protocol was reviewed and approved by the Institutional Review Board of the Johns Hopkins Bloomberg School of Public Health (Baltimore, MD) and the Ethics Committee of Asociacion Benefica PRISMA (Lima, Peru).

## Results

### Subject Characteristics

A description of the participants that were lost to follow-up and those that were retained is shown in Table 1. 45 percent of those retained for follow-up were male. The average HAZ was –1.99 and 53 percent were stunted at 24 months. The average years of maternal education was 8.04 years and the average per capita monthly income was $31.52. The average age at puberty was 12.64 among males and 11.79 among females in the group retained for follow-up. There was no significant difference in the gender breakdown, HAZ, percentage stunted at 24 months, years of maternal education or per capita monthly income in the children lost to follow-up.

**Table 1:**
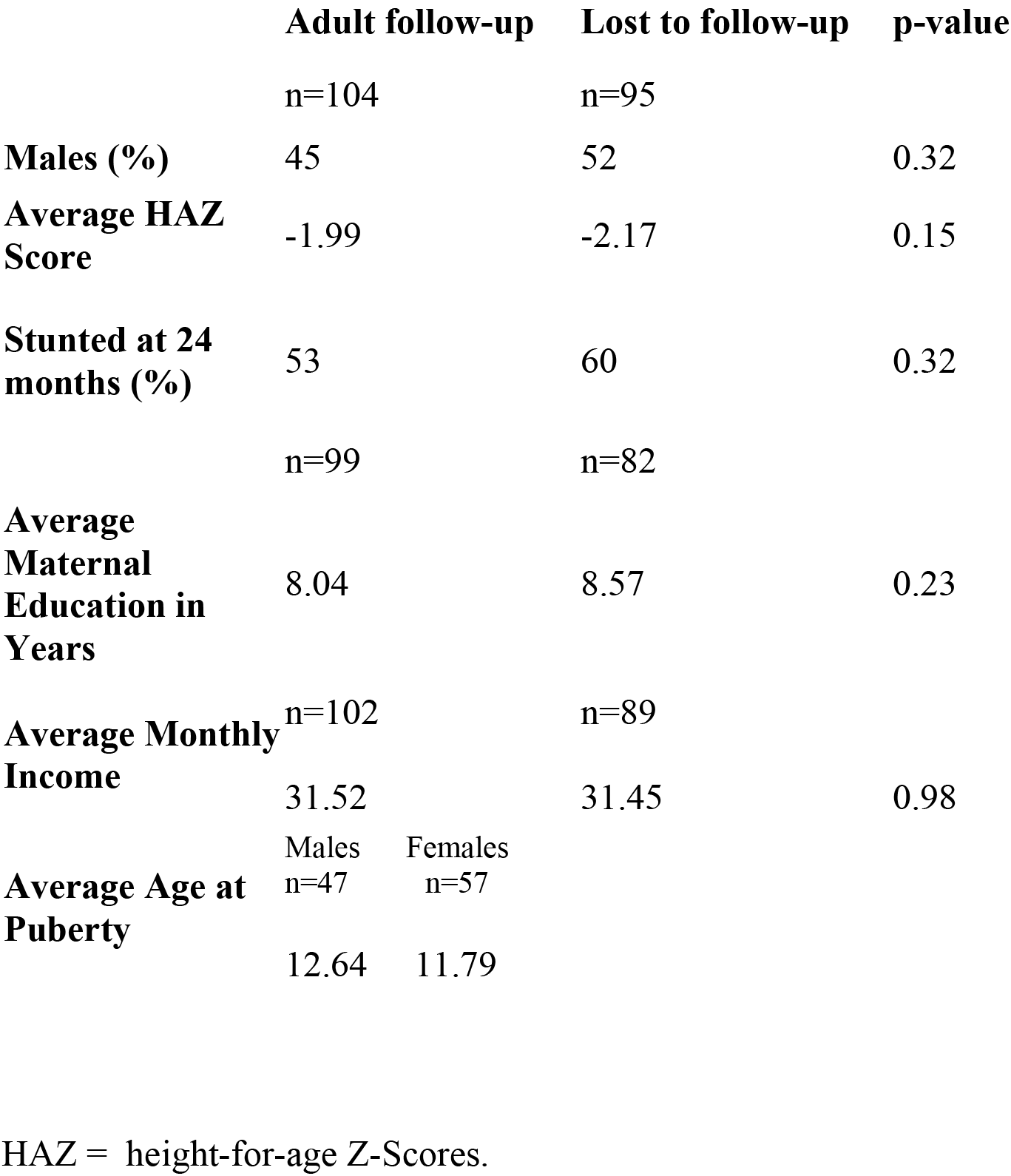
Comparison of participants with follow-up and those lost to follow up by gender, average HAZ scores, percent stunted at 24 months, average maternal education in years and per capita monthly income.

### Association of HAZ at 24 Months on HAZ, Weight, and BMI at Puberty

The mean HAZ, weight and BMI at puberty are presented in Table 2 categorized by participants who had a HAZ less than –2, HAZ between –2 and –1, and HAZ greater than –1 at 24 months.

**Table 2:**
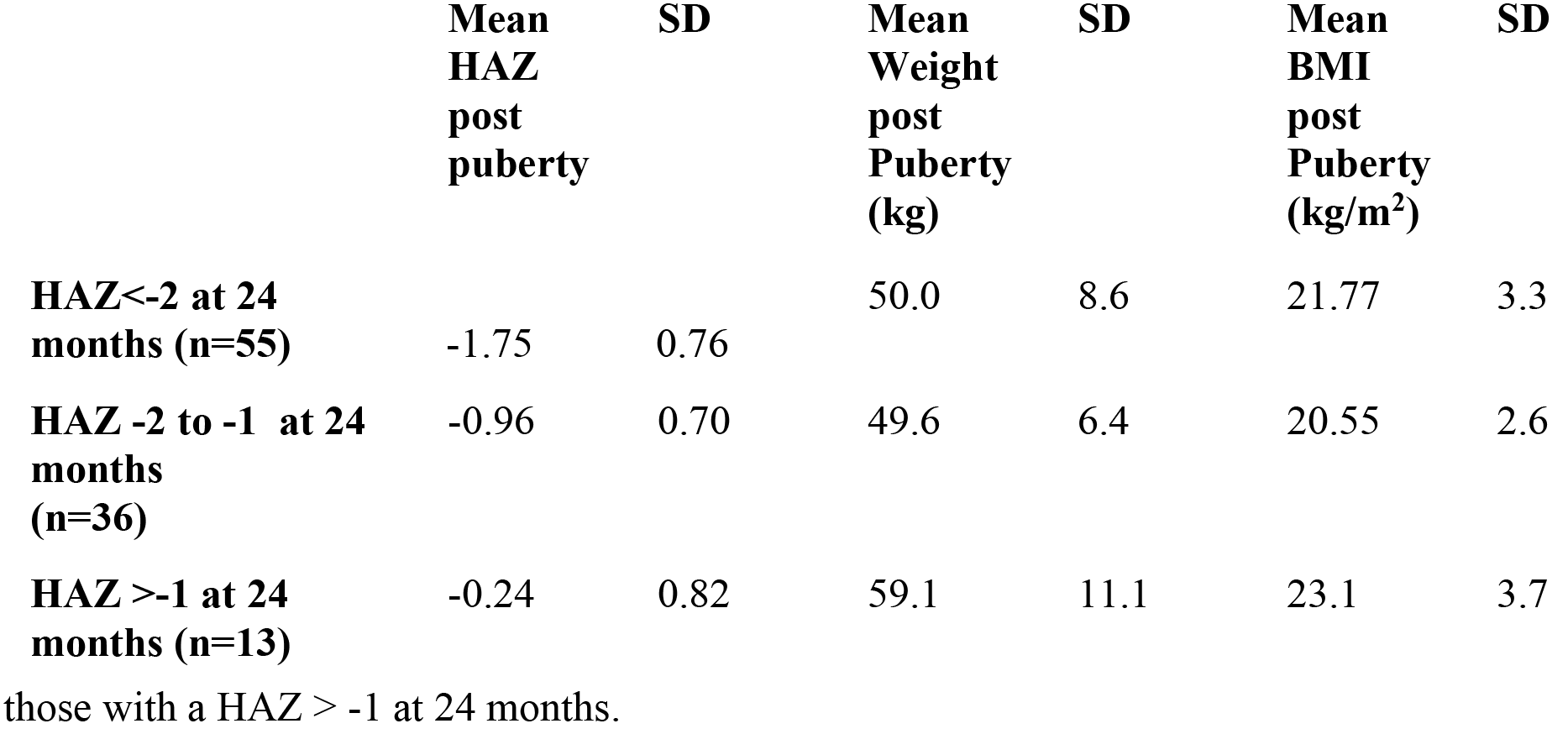
The mean HAZ at puberty, mean weight at puberty and mean BMI at puberty is shown by categorization of children who had a HAZ<-2, those who had a HAZ between –2 and BMI = Body Mass Index. Puberty was assessed by maternal report of the onset of menses (for female subjects) or the age of voice change (for male subjects).

When analyzing the association of HAZ at 24 months on HAZ at puberty there was a significant difference between groups as determined by one-way ANOVA (F(2,101) = 26.944, p <.001). A Tukey post hoc test revealed that the HAZ score at puberty was significantly lower if the child had a HAZ less than –2 at 24 months (−1.75±0.76, p<.001) and if the child has a HAZ between –2 and –1 at 24 months (−0.96±0.70, p =.011) compared to those who HAZ greater than –1 at 24 months (−0.24±0.82). The HAZ score at puberty was also significantly lower if the child had a HAZ less than –2 at 24 months (−1.75±0.76, p<.001) compared to those who had a HAZ between –2 and –1 at 24 months (−0.96±0.70).

The association of HAZ at 24 months on weight at puberty also showed a significant difference between groups as determined by one-way ANOVA (F(2,101) = 7.127, p =.001). A Tukey post hoc test revealed that the weight at puberty was significantly lower if the child had a HAZ less than –2 at 24 months (50.0±8.6, p =.002) and if the child had a HAZ between –2 and –1 at 24 months (49.6±6.4, p =.002) compared to those who HAZ greater than –1 at 24 months (−59.1±11.1). There was no significant difference in weight at puberty between children who had a HAZ less than –2 at 24 months and those who had a HAZ between –2 and –1 at 24 months (p =.975).

The association of HAZ at 24 months on BMI at puberty also showed a significant difference between groups as determined by one-way ANOVA (F(2,101) = 3.608, p =.031). A Tukey post hoc test revealed that the BMI at puberty was significantly lower if the child had a HAZ between –2 and –1 at 24 months (20.55±2.6 p =.034) compared to those who HAZ greater than –1 at 24 months (23.1±3.7). There was no significant difference in BMI at puberty between children who had a HAZ less than –2 at 24 months and those who has a HAZ between –1 and –2 at 24 months (p =.167). Similarly, there was no significant difference in BMI at puberty between children who had a HAZ less than –2 at 24 months and those who has a HAZ greater than –1 at 24 months (p =.347). In the three groups the mean BMI ranged from 20.55 to 23.1.

### Comparing Age Specific Prevalence of Stunting and Recovery by Gender

The overall prevalence of stunting was significantly different between boys and girls with rates of 66% and 42% (p = 0.015), respectively at 24 months (Table 3). The frequency of recovery in boys and girls recovered from stunting at 24 months to post-puberty was similar with boys at 68% and girls at 54%. Although there were different rates of stunting at 24 months the prevalence of stunting post-puberty was 23% for both sexes. Recovery from HAZ < –2 at 24 months to HAZ > –1 post-puberty was also similar among boys (23%) and girls (8%) at 24 months.

**Table 3:**
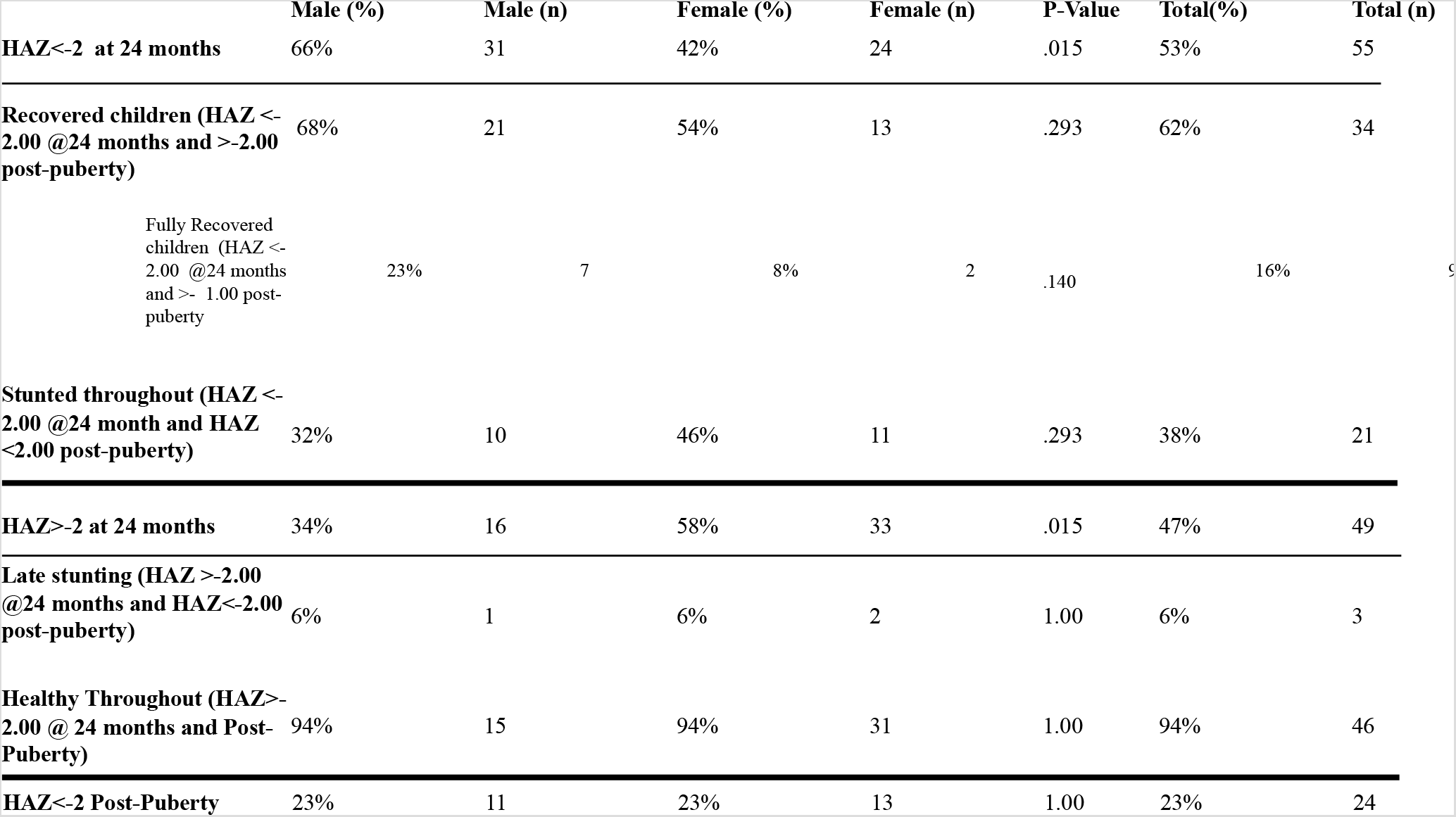
Prevalence of stunting and recovery from 24 months to post-puberty in males and females shown by categorization of children who recovered from stunting at 24 months, fully recovered from stunting at 24 months, those who remained stunted, those that became stunted in late childhood, and those that were neither stunted at 24 months or at their measurement following puberty.

The average age of puberty, as delineated by menarche in girls and voice change in boys is shown by categorization of children who recovered from stunting at 24 months, those who remained stunted, those that became stunted in late childhood, and those that were neither stunted at 24 months or at their measurement following puberty in Table 5. When analyzing the association of recovering from stunting on age at puberty in males there was a significant difference between groups as determined by one-way ANOVA (F(3,43) = 3.539, p =.022). A Tukey post hoc test revealed males who recovered from stunting had a significantly lower age of puberty (12.28±0.90, p =.007) compared to those who were stunted throughout (13.50±1.18). There was no significant difference in age of puberty between those who were stunted and those who were never stunted (p =.079) or between those who recovered from stunting and those who were never stunted (p =.621). When analyzing the association of recovering from stunting on age at puberty in females there was no significant difference between groups as determined by one-way ANOVA (F(3,53) =.139, p =.936).

### Rates of Recovery

In this cohort, it was found that a child who had a HAZ <-2 at 24 months was 9.5 times (CI 2.6–34.3, p-value 0.006) more likely to be stunted post-puberty compared to a non-stunted child at 24 months (Table 4). Although if a child is stunted at 24 months it is more likely that they will be stunted post puberty, the rates of recovery in this cohort were unexpectedly high. Figure 2 displays a matched comparison of attained length at 24 months and attained height after puberty demonstrates that in both boys and girls there is a trend towards recovery, with even those with severe stunting (HAZ≤-3) normalizing by the time they reached their adult stature. On the other hand, almost no children who were not stunted at 24 months became stunted later in childhood.

**Table 4:** Unadjusted odds ratios of stunted and non-stunted children who will be stunted post-puberty (PP)

|  | **OR** | **CI** | **n** | **P-Value** |
| --- | --- | --- | --- | --- |
| **Odds Male HAZ<-2 @ 24 m Stunted PP Compared to Non-Stunted Male @ 24 m** | 9.5 | 2.6, 34.3 | 47 | 0.006 |
| **Odds Female HAZ<-2 @ 24 m Stunted PP Compared to Non-Stunted Female @ 24 m** | 13.1 | 2.5, 67.6 | 57 | 0.002 |
| **Odds Child HAZ<-2 @ 24 m Stunted PP Compared to Non-Stunted Child @ 24 m** | 7.1 | .82, 61.9 | 104 | 0.074 |

**Table 5:**
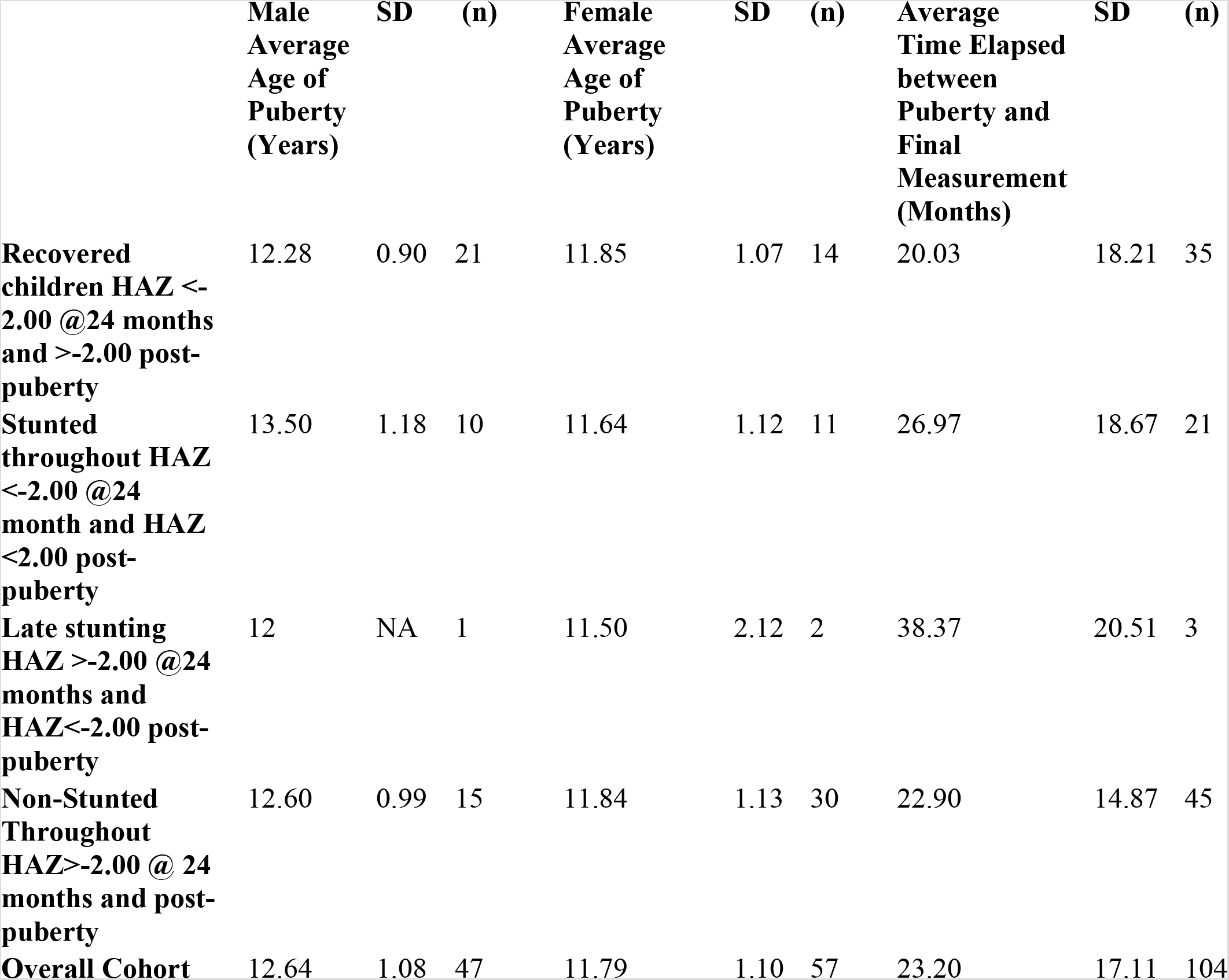
The average age of puberty, as delineated by menarche in girls and voice change in boys is shown by categorization of children who recovered from stunting at 24 months, those who remained stunted, those that became stunted in late childhood, and those that were neither stunted at 24 months or at their measurement following puberty.

|  | Male<br>Average<br>Age of<br>Puberty<br>(Years) | SD | (n) | Female<br>Average<br>Age of<br>Puberty<br>(Years) | SD | (n) | Average<br>Time Elapsed<br>between<br>Puberty and<br>Final<br>Measurement<br>(Months) | SD | (n) |
| --- | --- | --- | --- | --- | --- | --- | --- | --- | --- |
| **Recovered children HAZ <-2.00 @24 months and >-2.00 post-puberty** | 12.28 | 0.90 | 21 | 11.85 | 1.07 | 14 | 20.03 | 18.21 | 35 |
| **Stunted throughout HAZ <-2.00 @24 month and HAZ <2.00 post-puberty** | 13.50 | 1.18 | 10 | 11.64 | 1.12 | 11 | 26.97 | 18.67 | 21 |
| **Late stunting HAZ >-2.00 @24 months and HAZ<-2.00 post-puberty** | 12 | NA | 1 | 11.50 | 2.12 | 2 | 38.37 | 20.51 | 3 |
| **Non-Stunted Throughout HAZ>-2.00 @ 24 months and post-puberty** | 12.60 | 0.99 | 15 | 11.84 | 1.13 | 30 | 22.90 | 14.87 | 45 |
| **Overall Cohort** | 12.64 | 1.08 | 47 | 11.79 | 1.10 | 57 | 23.20 | 17.11 | 104 |

**Figure 2:**
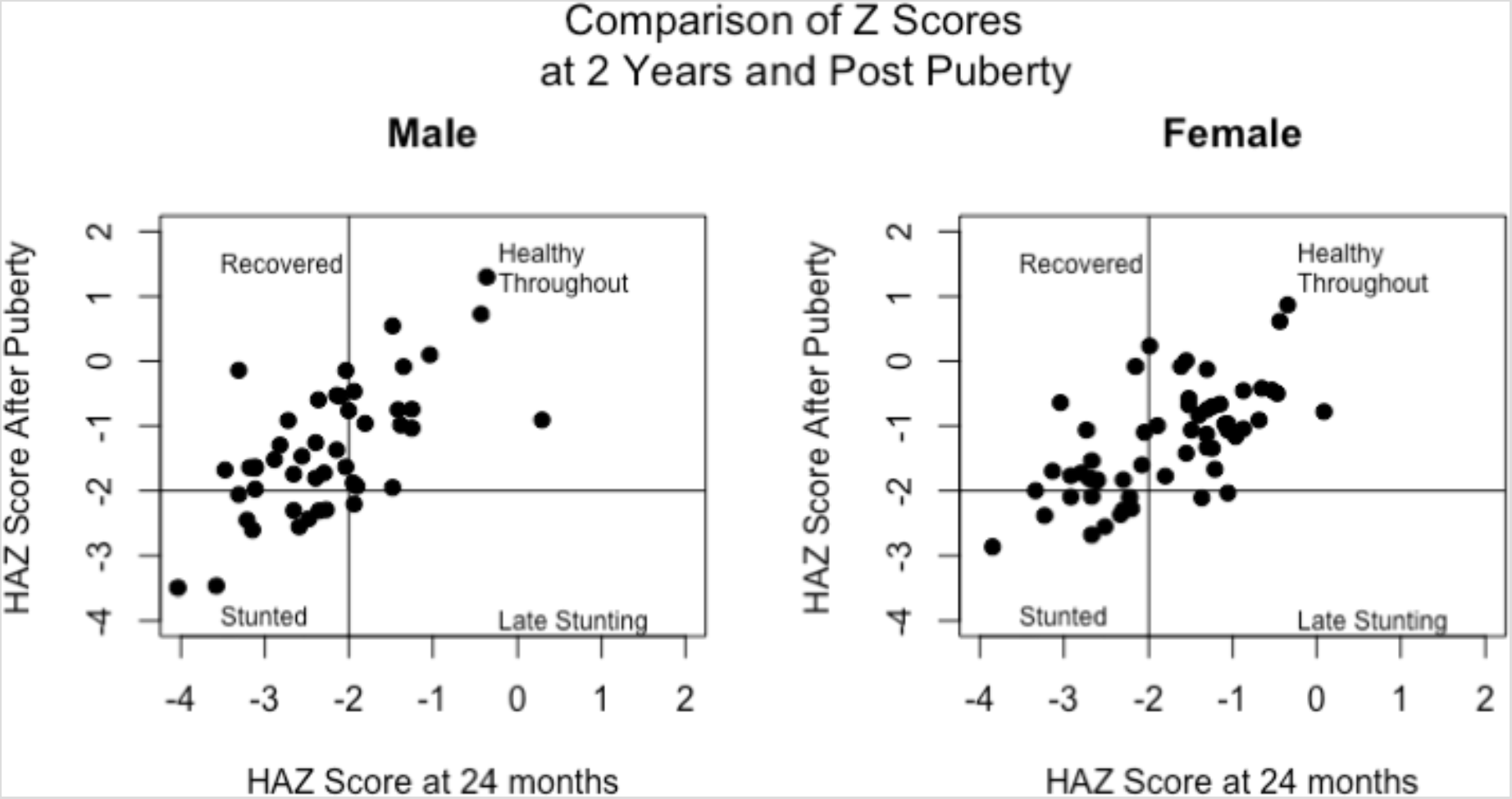
A matched comparison of attained length at 24 months and attained height after puberty. X- axis shows HAZ score at 24 months and Y axis shows the corresponding HAZ score after puberty. Each point represents a single participant. The graph is broken into four quadrants to delineate children who at puberty had recovered from stunting at 24 months, those that were neither stunted at 24 months or at their measurement post-puberty, those that became stunted in late childhood, and those who remained stunted.

Among the birth cohort in Santa Clara, there was recovery from stunting among both girls and boys, with 62% (n = 34) of children who had a HAZ <-2 at 24 months (n = 55) achieving a HAZ > –2 post-puberty (Table 3). More specifically, 16% (n = 9) of children who were stunted at 24 months (n = 55) had a HAZ>-1 post-puberty.

Despite high rates of recovery from stunting, the mean attained height among cohort members is well below the expected attained height as reported by the WHO. Among the male cohort members, the mean anthropometrical measurements post puberty were a height of was 156.38 cm (SD 6.34), HAZ score of –1.34 (SD 1.01), weight of 52.15 kg (SD 9.79) and BMI of 21.26 (SD 3.27). Among the female cohort members the mean anthropometrical measurements post puberty were a height of 151.69 (SD 5.74), HAZ score of –1.24 (SD 0.84), weight of 50.00 (SD 7.78) and BMI of 21.73 (SD 3.16). This result indicated that the average man in Iquitos is 11.31 cm shorter and the average woman in Iquitos is 9.20 cm shorter than mean attained adult standard as indicated by WHO 2006 norms.

## Discussion

Our results indicate significantly higher rate of stunting in boys than girls at 24 months. After puberty, boys still had slightly higher but not statistically significant rate of stunting. Although being stunted at 24 months of age was a risk factor for having a constrained adult height, the majority of children who were stunted at 24 months exhibited growth recovery, including children with HAZ of ≤-3 at 24 months. If children were not stunted by 24 months of age, they were unlikely to become stunted in later childhood. Our interpretation of this data is that the potential for linear growth recovery in childhood does exist. The use of the HAZ at 24 months is a useful one to target populations of children who would may benefit from intensified dietary and health interventions to maximize statural growth in late childhood. On the individual level, parents of children who are stunted at 24 months should be encouraged to optimize current diet to promote growth in later childhood, which may allow for normal adult stature to be attained.

A weakness in the present study was the loss to follow-up, which was 48%. Nearly all this loss was due to migration outside the study area. Of the group that was not able to be assessed for the ascertainment of puberty, 48% had a known migratory destination, which was most likely to be Lima. When examined, there was no significant difference in the HAZ at 24 months and eventual migratory status. Even if located the change in environment would have limited our ability to interpret the data, loss to follow-up is difficult to avoid with a gap in time to follow-up of 12–14 years.

This study is novel in showing that higher rates of stunting among boys than girls in early childhood dissipate after puberty. This brings up the question of when the crucial catch-up growth occurs in boys. It has been suggested that late childhood presents another critical window when interventions can be made to increase height growth.^21^ A calcium carbonate supplement given for 13 months to British boys age 16–18 years was associated with a modest increase of 7 mm compared to the control group.^22^ Additionally, a 12 month prepubertal calcium carbonate supplementation given to Gambian boys age 8–12 years increased heights two centimeters at age 15.5 in comparison to the control group. Although the calcium group had a greater stature in adolescence, subsequently, the calcium group stopped growing earlier and was 3.5 at a mean age of 23.5 y. This population commonly has late onset puberty and low calcium intake.^23^ This seems to suggest that interventions in late adolescence may be more effective in males, but as with most nutritional supplementation studies, the absolute amount of the total attained height deficit corrected by the intervention is quite limited. Clearly though, more work needs to be done to study this window for catch-up growth. Tumilowicz et al. advocates specifically for future nutrition research that explores standard indicators to measure adolescent dietary quality to determine causal effects between dietary intake and growth.^24^

Among boys and girls, this nonintervention-study showed high rates of recovery with 62% of children in the birth cohort who had a HAZ<-2 at 24 months having a HAZ>-2 post-puberty, and 16% of children stunted at 24 months fully recovering with HAZ>-1 post-puberty. These rates of recovery suggest that catch-up growth after 24 months is possible and common among children who stay in resource constrained environments with a high prevalence of acquired linear growth deficits. Our study adds to a large amount of literature that suggests there is a natural high rate of recovery from stunting after the first thousand days. Walker et al. found 87% of participants who received a nutritional intervention from 24–48 months were no longer stunted at follow-up at 7 to 8 years. This rate of recovery, while promising, did not differ from the control, a group of children who were stunted at 24 months and not given any intervention.^25^ Satarayana et al. found that Indian girls stunted in childhood gained significantly more height in the years before finishing puberty than girls who were not stunted in childhood, even with no nutritional intervention.^26^ The Young Lives study in Peru-which includes participants from urban and rural areas and represents coastal, highland, and jungle regions-found that one-third of children were stunted around 1 year of age had recovered (HAZ>-2) at age 5, again with no nutritional intervention.^27^

The adolescent growth spurt has been posited as a critical period of growth beyond the first 1000 days.^21^ This may explain why our study, in which we did follow-up measurements after puberty, showed much higher rates of recovery from stunting than other studies. The Young Lives study in Peru, which includes participants from urban and rural areas and represents coastal, highland, and jungle regions, found that one-third of children were stunted around 1 year of age had recovered (HAZ>-2) at age 5.^27^ Sterling et al. found among 27 participants that were stunted at 12 months only 11 recovered by adolescence (40%).^28^ In contrast, we found that of 55 participants who were stunted at 24 months, 34 had recovered post puberty (62%) despite higher rates of stunting in early childhood as would be expected given the location of the study site in one of the poorest regions of Peru. The major difference between these studies and ours is that follow-up in our study was done only after participants went through puberty. This suggests that there is significant room for recovery from stunting through adolescence, even in conditions where stunting is common and severe.

Puberty is a time of significant growth among girls and boys and it is key to look at the onset of the end of puberty when looking at catch-up growth. Our study found the mean age of menarche to be 11.8 (median 12) and the mean age of voice change to be 12.6 (median 12), whereas the median age at menarche was found to be 12.8 years and the median for voice change 15.1 years among adolescents in Germany.^29^ The only group that had a significantly later onset of puberty was boys who were stunted both at 24 months and post puberty. The early onset of puberty in this population is another signal of increased nutrition, as in many other populations with high percentages of early childhood stunting, menarche and voice change are significantly delayed.^30, 31, 32^ The early onset of puberty may be one reason that although significant recovery was made from stunting, the average height attained by the cohort was still more than one standard deviation below the WHO standards. Studies following malnourished children adopted to more developed countries have found that early puberty can negate the gains made in childhood.^33^ This is of particular attention as obesity is an ever-growing problem even in developing countries. Within this cohort all categories of children had a healthy mean BMI. Many think over-nutrition can lead to early puberty, meaning that nutritional interventions must be carefully timed and carried out both to avoid obesity and extend the pubertal growth spurt.^34^

One of the most damaging aspects of growth stunting is its association with delayed cognitive development in childhood, reduced height and lower earning potential as an adult.^6^ There has been controversy over whether or not recovery in stature from growth stunting corresponds to recovery from the cognitive effects of stunting. The majority of the literature, however, now seems to agree that those who recover from stunting have similar cognitive ability to those who were never stunted.^27, 35, 36, 37^

In addition to the risk of delayed cognitive development, stunted maternal height causes a multigenerational burden. An analysis of 109 Demographic and Health Surveys in 54 countries conducted between 1991 and 2008 found that a 1-cm increase in maternal height was associated with a decreased risk in underweight (RR, 0.968; 95% CI, 0.968–0.969), stunting (RR, 0.968; 95% CI, 0.967–0.968), and wasting (RR, 0.994; 95% CI, 0.993–0.995).^38^ Maternal stunting increases the risk of giving birth to children who are small-for-gestational age (SGA). This is of particular concern because multiple studies show that birth size affects development^39^. Additionally, Black et al. estimated that one-fifth of all stunting originates in the fetal period, resulting from children born SGA, which perpetuates the cycle of stunting that occurs in many LMICs. Nutritional interventions focused on adolescent women can reduce risks of complications in pregnancy and improve the fetal growth and development for their future children. Providing iron-rich school meals has been shown to reduce iron-deficiency anemia, which increases risk maternal mortality, low birthweight, and perinatal mortality.^39^

Multiple micronutrient interventions are another strategy that has been successful in improving height and weight gains among both male and female children. These studies, which ranged in length, provided the daily-recommended allowance of iron, zinc, vitamin A, folic acid, and B vitamins to children ranging from 6 months to 11 years in Latin America, Africa, and Asia.^40^ While effective, multiple micronutrient are difficult to carry out on a large scale because of high costs. Nutritional-sensitive interventions, or programs that address the underlying causes of malnutrition and improve the effects of nutrition-specific interventions, may be a more effective long-term strategy to improve nutrition throughout a lifecycle. Biofortification of crops is one promising strategy to help provide a daily dose of micronutrients and prevent deficiencies.^41^ Further research is necessary to evaluate the impact of biofortification on stunting. Continued promotion of schooling, for boys and girls, is also an important strategy for the nutritional health of the next generation. Maternal schooling level has been positively associated with child nutritional status.^42^

This study adds to the growing literature that recovery from stunting is possible and common and that the predictive capacity of growth at two years, while an important risk indicator for poor outcomes, is not as tightly associated with adult height as is commonly held. More research needs to be done to identify what promotes catch-up linear growth in late childhood. Some studies have suggested that the major predictor of adult stature is the severity of stunting in infancy, though this study did not find that.^27^ The first 2 years of life are crucial for growth and development, and efforts should continue to make sure that every child receives proper nutrition and care during this critical time. However, a child who is stunted at 24 months should not be viewed as stunted- in stature or cognition-for life. Throughout childhood and adolescence nutritional programs have value to allow stunted children to achieve full genetic height potential by the time they become adults, and to reduce stunting and associated morbidities in future generations.

## Data Availability

Data is available from the corresponding author upon request.

## List of Authors Information

Julia Schwarz, Icahn School of Medicine 1 Gustave L. Levy Pl, New York, NY 10029;

Pablo Peñataro Yori, Johns Hopkins Bloomberg School of Public 615 N. Wolfe Street Room E5532 Baltimore, Maryland 21205;

William K. Pan, Duke University 227 Trent Hall, 310 Trent Drive, Durham, NC 27708;

Maribel Paredes Olortegui, Asociación Benéfica Prisma Calle Ramirez Hurtado, 622 Iquitos – Perú;

Robert Klapheke, UC San Diego School of Medicine 9500 Gilman Drive La Joalla, CA 92093;

Margaret N. Kosek, Johns Hopkins Bloomberg School of Public Health 615 N Wolfe St Baltimore, MD 21205;

